# Joint estimation of the effective reproduction number and daily incidence in the presence of aggregated and missing data

**DOI:** 10.1101/2024.06.06.24308584

**Authors:** Eamon Conway, Ivo Mueller

## Abstract

Disease surveillance is an integral component of government policy, allowing public health professionals to monitor transmission of infectious diseases and appropriately apply interventions. To aid with surveillance efforts, there has been extensive development of mathematical models to help inform policy decisions, However, these mathematical models rely upon data streams that are expensive and often only practical for high income countries. With a growing focus on equitable public health tools there is a dire need for development of mathematical models that are equipped to handle the data stream challenges prevalent in low and middle income countries, where data is often incomplete and subject to aggregation. To address this need, we develop a mathematical model for the joint estimation of the effective reproduction number and daily incidence of an infectious disease using incomplete and aggregated data. Our investigation demonstrates that this novel mathematical model is robust across a variety of reduced data streams, making it suitable for application in diverse regions.

**Author summary:** Monitoring the transmission of infectious diseases is an important part of government policy that is often hindered by limitations in data streams. This is especially true in low and middle income countries where health sectors have less funding. In this work we develop a mathematical model to enhance disease surveillance by overcoming these data limitations, providing accurate inferences of relevant epidemiological parameters.

## Introduction

During the COVID-19 pandemic, effective and timely infectious disease surveillance was an integral part of government policy [1]. This emphasis on disease surveillance led to the development and extension of tools that can provide efficient and reliable updates on the effective reproduction number [2–6]. However, the majority of these tools rely upon a line list of infections or regular daily measurements of case incidence, measurements that can only be obtained with consistent and dedicated effort by the countries’ government and health care sectors.

The cost of daily surveillance during the pandemic was deemed warranted, but as COVID-19 has become widespread, endemic and a part of daily life, the majority of countries have reduced their surveillance effort and are resorting to reporting weekly (or larger) windows of case data. For example, the public reporting of COVID-19 infections within Australia is now weekly, in line with the reporting of other infections such as influenza [7]. This change to reporting timelines leads to a reduced load on the health sector, but does come with the cost of reducing the fidelity of data, on which mathematical models rely [1, 8]. To understand this reduced data landscape new mathematical methods must be developed.

In recent work [4, 9], it is highlighted that there is a need for algorithms that can calculate the effective reproduction number from aggregated data. To address this need an expectation-maximisation algorithm has been proposed for use within the established 4, EpiEstim framework [ 10]. The algorithm maximises the probability of the daily incidence given the aggregated data by treating the effective reproduction number as a latent variable. In doing so, a maximum likelihood approximation for the daily incidence is obtained, which is then used with EpiEstim’s standard methods to calculate the effective reproduction number.

The cost of disease surveillance in countries that have more limited health systems capacity is often overlooked during the development of novel surveillance techniques. In these regions, the money available to be spent on surveillance is small and the data on case counts may be subject to inconsistent reporting and drop out. This irregular data can limit the application of modern mathematical models, even in the best case scenario when dropout occurs for only a short period of time. For example, [11] state that fourteen days of uninterrupted measurements are required to get accurate estimates from EpiNow2. This requirement imposes a critical constraint on the health systems for accurate measurement of the effective reproduction number, a requirement that some regions are not equipped to handle. As such, obtaining realistic measurements of the effective reproduction number in the presence of data dropout is an integral requirement of modern epidemic analysis.

The number of epidemic analysis tools that are developed with the explicit inclusion of data dropout is limited. As such, people often resort to *a-priori* data imputation techniques, filling in any missing data to allow the application of existing inference techniques. This may be problematic as the inference techniques treat the imputed data as if it was obtained from the same observation model as the true data. This assumption reduces the true uncertainty that is present in the data and produces biases dependent upon the imputation technique [12–14], possibly resulting in misleading and overly certain estimates.

In this work we develop a novel mathematical model for the inference of both the effective reproduction number and daily incidence from data streams with data dropout and aggregated measurements of cases. Importantly, our model retains the ability to make effective inferences when working with incomplete data. Furthermore, due to the generality of the infection process our model is applicable to inferring the effective reproduction number in other notifiable infectious diseases and not just COVID-19. Our novel mathematical model is an invaluable tool for infectious disease surveillance, especially when surveillance is sporadic or has become too costly.

## Methods

We develop a Bayesian method to estimate the effective reproduction number and daily incidence from *N* observed aggregate case counts over known time intervals. To do so,

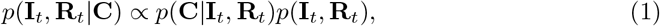

we define an expression for, where **I**_*t*_ = [*I*_1_, *I*_2_, …, *I*_*t*−1_, *I*_*t*_] and **R**_*t*_ = [*R*_1_, *R*_2_, …, *R*_*t−*1_, *R*_*t*_] are the vectors of random variables containing the daily incidence and effective reproduction number up to and including day *t* respectively, and **C** = [*C*_1_, *C*_2_, …, *C*_*N*_] is the vector containing the measured case counts.

We state that the *i*-th measurement of cases can be described by,

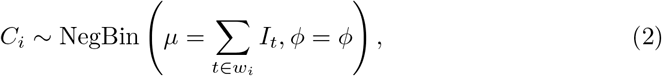

where NegBin denotes the negative binomial distribution parameterised by mean *µ* and shape *ϕ*^1^, *I*_*t*_ is the incidence on day *t* and *w*_*i*_ is the known window of days over which the *C*_*i*_ cases were measured. By assuming that each observation of cases is conditionally independent,

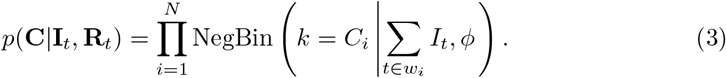

In contrast to other work, where the daily incidence is treated as a known quantity, we allow each *I*_*t*_ to be a random variable that is to be inferred from our observed data. For our purposes we assume that observations of **I**_*t*_ follow a simple and widely used mathematical model for disease transmission.

To define, *p*(**R**_*t*_, **I**_*t*_) = *p*(**I**_*t*_|**R**_*t*_)*p*(**R**_*t*_), we use an infectious disease model such that,

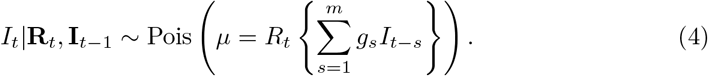

Here *g*_*s*_ is the density of the generation interval on day *s* and *m* is the last day of infectivity. Therefore a suitable prior for daily infections given the effective reproduction number is,

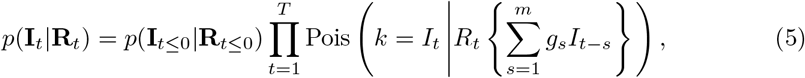

where *T* = max(*w*_*N*_) is the largest day of interest and *p*(**I**_*t≤*0_|**R**_*t≤*0_) is the prior information required for estimating daily incidence before the time window of interest.

To define a prior on daily reproduction number we assume that **R**_*t*_ follows a random walk model. Therefore,

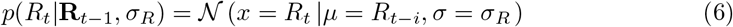

where 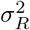 is the variance of the random walk. This gives,

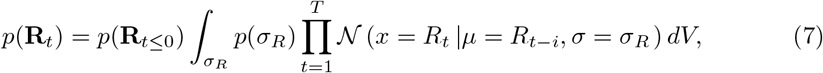

where *p*(**R**_*t≤*0_) is probability distribution for the effective reproduction number before our time window of interest. Finally, we assume that,

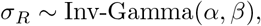

and

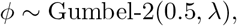

where we have used a Type-2 Gumbel distribution as an approximation of the penalised complexity prior for the shape parameter in the negative binomial distribution [15, 16] and *α*, *β*, and *λ* are known constants encoding our prior knowledge (for the choice of *λ* see Appendix C). By combining Equation 3, Equation 5 and Equation 7 into Equation 1, we arrive at the distribution,

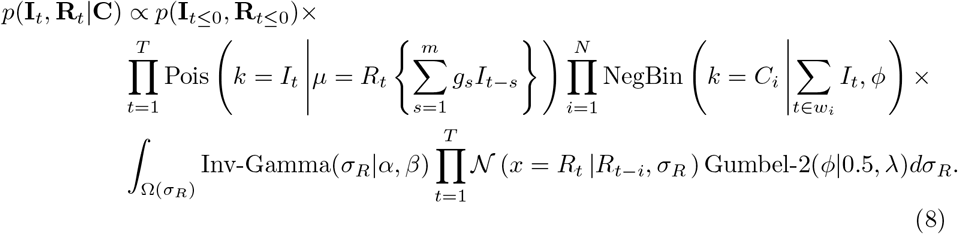

It is important to note that we do not make any requirements on the time windows over which measurements are made, allowing the model to also estimate any *I*_*t*_ and *R*_*t*_ that do not belong to a time window, as well as naturally handling overlapping windows.

## Initial conditions

Within our posterior, Equation 8 there are values required for both incidence and effective reproduction numbers before the window of inference. This occurs for *R*_1_ and for *I*_1_ to *I*_*m*_. Therefore we must provide reasonable estimates for any *R*_*t*_ and *I*_*t*_ required outside of our window of inference, however we cannot continue with the recursive relationships that were used previously.

We argue that the prior for the initial effective reproduction number should be distributed around one,

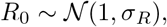

and that the prior for the daily incidence is constant,

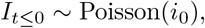

where *i*_0_ a known constant estimate on the starting cases. This leads us to the conclusion that the most appropriate choice is,

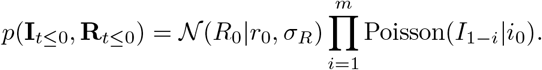

We note that in the case where daily data can be obtained prior to the aggregated windows, we can use that daily information to more accurately inform our initial conditions.

## Numerical implementation

We use Stan to generate samples from the posterior distribution [17], however there is a requirement that the sampled random variables are continuous. Therefore, we assume that **I**_*t*_ is a continuous random variables and that,

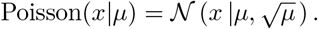

We expect that this Normal approximation for the Poisson distribution will be accurate for all regimes of interest as typically the value for *µ* is large. All code used to generate results is available at https://github.com/EamonConway/reduced_data_model.

## Results

To validate our novel mathematical model we first perform inference on a simulated data set with known daily values for the effective reproduction number and incidence. We then present results for real data observed in Victoria, Australia during the COVID-19 pandemic [18].

### Simulated data

For our simulated data set, we assume that

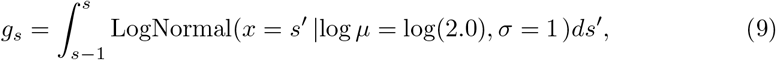

and

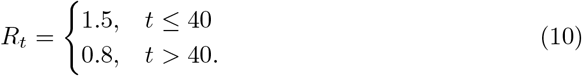

Simulated data for the daily incidence is then generated by the recursive relationship,

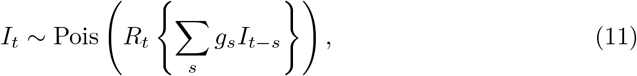

where *I*_0_ = 100 and *I*_*t*_ = 0 for t ∈(∞, 1].

The first example that we consider consists of aggregating our known simulated daily data into seven day windows. In Figure 1 we demonstrate the joint estimation of both the effective reproduction number and the daily incidence. Figure 1a shows great agreement between the estimates for the effective reproduction number and the known *R*_*t*_, but with an obvious smoothing over the shock at *t* = 40. The smoothing over the shock in the effective reproduction number is due to the size of the aggregation window, as the aggregated data does not provide enough information for the inference algorithm to accurately capture the exact moment that the effective reproduction number is changed. We can see that in Figure 1b the algorithm has obtained an accurate estimate for the daily incidence except for around the shock. The smoothing of the shock has resulted in the peak incidence being outside of the 95% quantiles. To demonstrate the effect of the aggregation window on inferring shocks in effective reproduction number, we present results in Appendix A.1 for differing window sizes. It is obvious that as the aggregation window is decreased, we more accurately capture the true behaviour of the simulated data.

**Fig 1.**
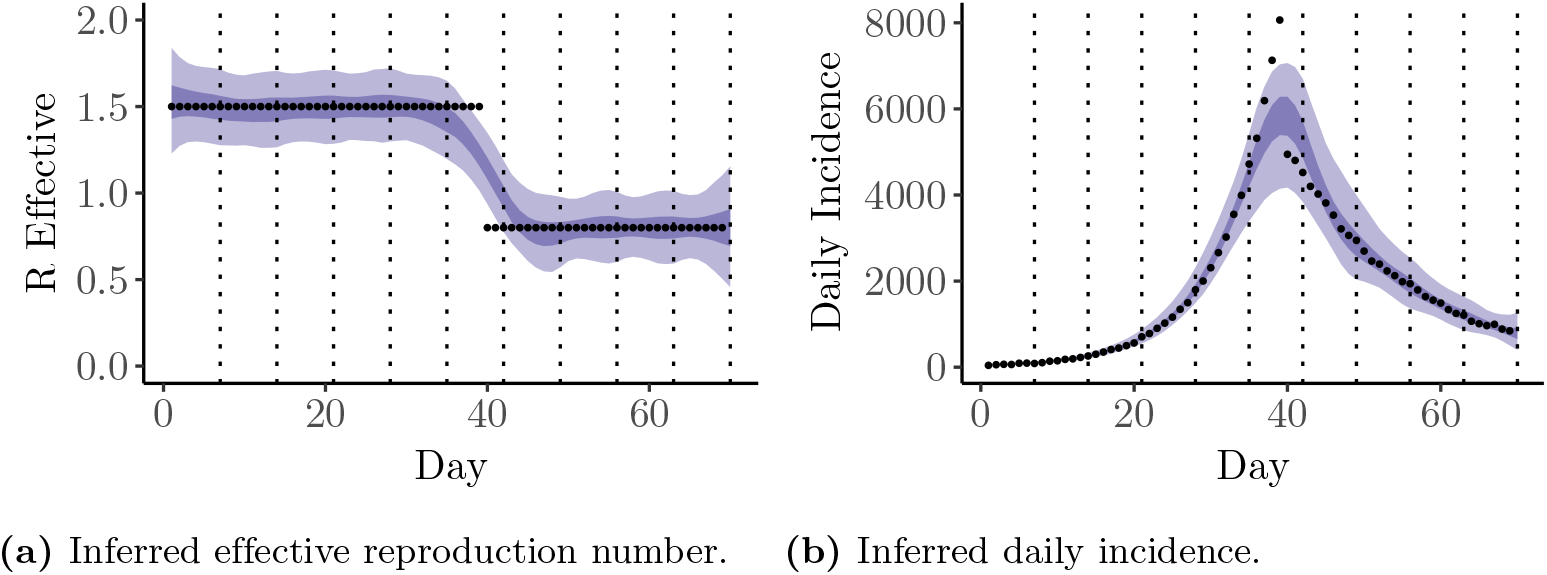
Application of the proposed inference method to seven day aggregated simulated data where *α* = *β* = 10^−3^ and *λ* = log 2. The ribbons correspond to the 50% and 95% quantiles. The dashed lines are indicate the aggregation windows.

To test the validity of our inference method in the presence of missing data, we remove 50% of our simulated observations at random, resulting in quite a severe level of dropout. In Figure 2 we see that even with this minimal data we can obtain reasonable estimates for the effective reproduction number and epidemic trajectory. We observe that that there is less smoothing in the effective reproduction number, Figure 2a, in comparison to the aggregated data set. In direct contrast to the aggregated data, the dropout data set is able to more accurately determine the location of the shock in the effective reproduction number as there are consistent observations in the regions of the shock. We also note that the quantiles for the inferred daily incidence, Figure 2b, tightly follow the known simulated incidence.

**Fig 2.**
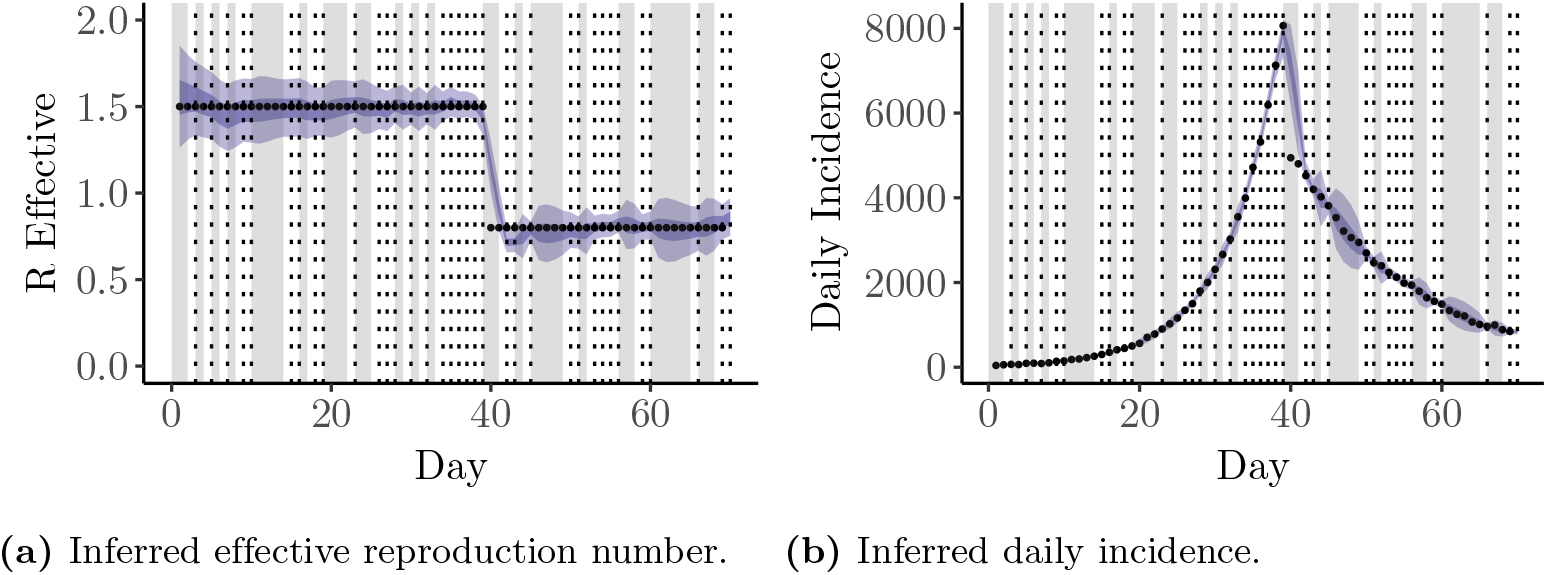
Application of the proposed inference method to simulated data with 50% random dropout, where *α* = *β* = 10^−3^ and *λ* = log 2. The ribbons correspond to the 50% and 95% quantiles. The dashed lines indicate the measurement times and the grey area is missing data.

To further support the validity of our inference method we provide many more worked examples with simulated data in Appendix A, including examples with both aggregation and dropout together. Importantly, in all examples we recover reasonable estimates for the effective reproduction number and the daily incidence. This highlights the robustness of our inference method and assures us that the technique is valid for application on real-world data.

### Real Data

Having validated our novel inference algorithm on simulated data, we now test its use on known case data from the COVID-19 pandemic in Melbourne, Victoria for 105 days from 1st September 2021 [18]. For this work we use the generation interval reported in [11], which is a refitting of the work in [19] using the incubation period reported in [20].

There is no definitive measurement of the true effective reproduction number when working with real world data. Therefore, we determine a daily effective reproduction number from the posterior,

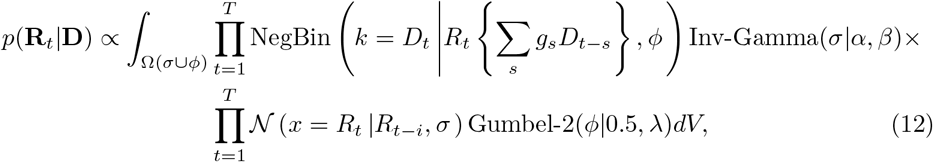

where **D** is the vector of observed daily incidence and *D*_*t*_ is the daily observation of incidence at time *t*. Equation 12 is the direct analogue of Equation 8 when the daily incidence is not treated as a random variable and there is no aggregation or dropout. We note that this equation is similar, albeit with different assumptions for the priors and the time varying nature of the effective reproduction number, to the posterior used in EpiNow2 [2]. For ease of reference we will denote Equation 8 as the reduced data model and Equation 12 as the daily data model.

We investigate the applicability of our inference algorithm by aggregating the known real-world daily case data into seven day windows. This is reflective of the current reporting status in Australia and other countries, where COVID-19 figures are aggregated weekly. Figure 3 depicts the inferred effective reproduction number and daily incidence from the aggregated data. We observe that the reduced data model adequately captures all of the general trends in the effective reproduction number when compared to the daily model (Figure 3a). Furthermore, our joint estimation technique has allowed for the recovery of the daily incidence as well, this is shown in Figure 3b, where we can see good agreement between our estimates and the known daily observations of incidence.

**Fig 3.**
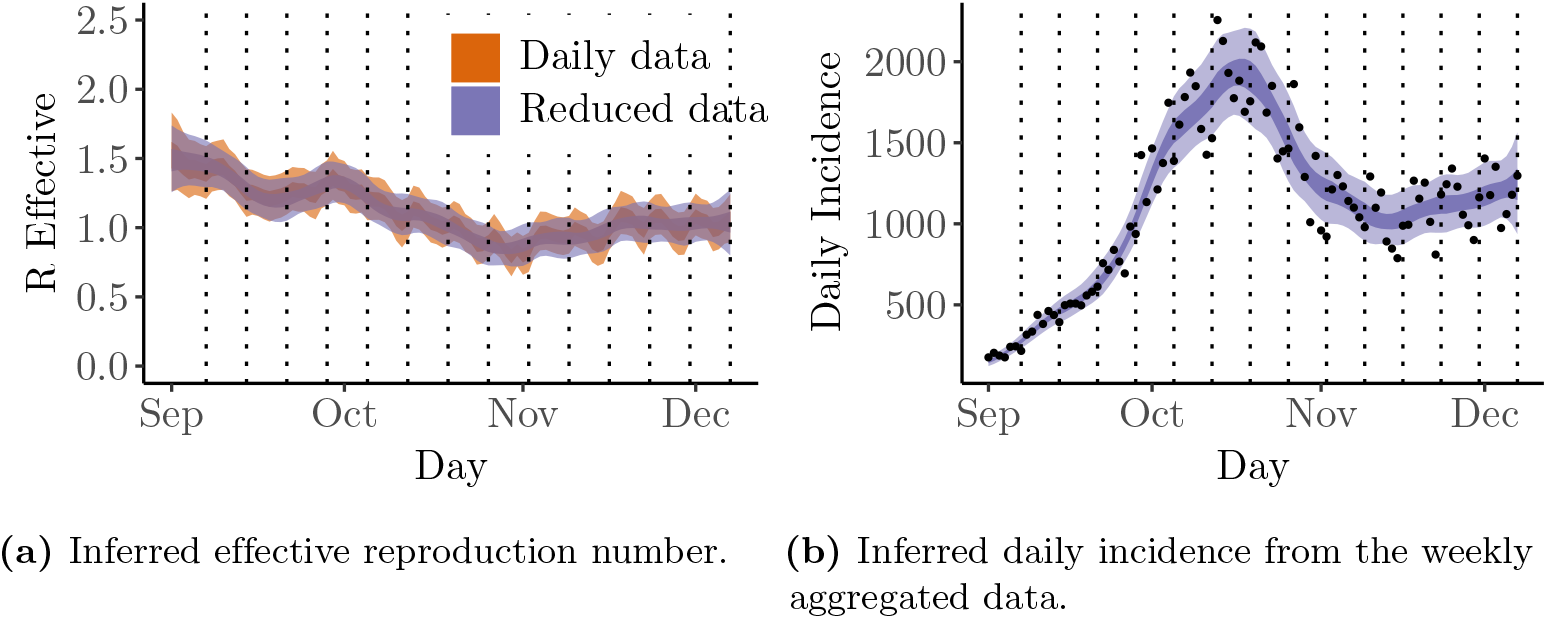
Comparison of the reduced data model using seven day aggregated real data from the COVID-19 pandemic in Victoria, Melbourne and the daily model. Here, *α* = *β* = 10^−3^ and *λ* = log 2. The ribbons correspond to the 50% and 95% quantiles and the dashed lines are the day of reporting.

Surveillance techniques used to infer the effective reproduction number should be insensitive to unforeseen changes in their data stream. This is especially important at the start of a pandemic where new data streams are established, a complicated process which may result in data dropout. To demonstrate how the reduced data model performs in the presence of arbitrary data dropout, we construct a low fidelity data set by randomly sampling 51 data points from the observed real data. Presented in Figure 4 are the results of applying our novel technique to the dropout data set. By a direct comparison to the daily data model, Figure 4a, we can see that we have adequately captured the effective reproduction number and successfully obtained estimates for the missing daily incidence. It is obvious from the quantiles that there is more uncertainty in regions which are missing larger amounts of data. To exemplify this behaviour, we highlight the time period in November where there is a large amount of missing data in our low fidelity data set. This time period corresponds to increased quantiles in both effective reproduction number and daily incidence. However, even with this level of dropout the reduced data model is able to provide reasonable estimates for what happened during and immediately after the period of dropout.

**Fig 4.**
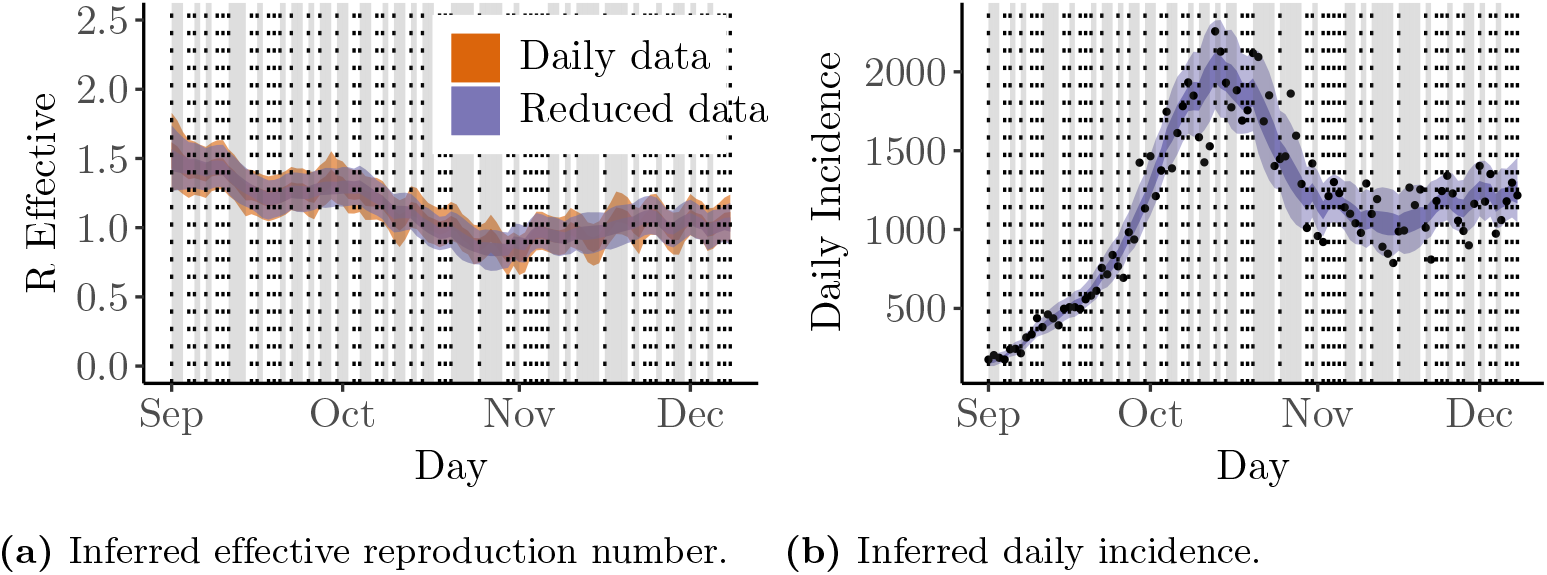
Inferring the effective reproduction number and daily incidence from 51 randomly chosen days of real data from the COVID-19 pandemic in Melbourne, Victoria. Here, *α* = *β* = 10^−3^ and *λ* = log 2. The ribbons correspond to the 50% and 95% quantiles. The dashed lines are the day of reporting and the grey areas are regions of missing data.

We highlight that the reduced data model is also robust to the dropout of data on the aggregated level for real data. We provide results for windows of varying sizes with 50% random dropout within Appendix B. The reduced data model performs better with more data and smaller windows of aggregation, but is still flexible enough to overcome data dropout even with aggregation windows.

As priorities evolve within the health sector it is feasible that the window of aggregation will vary. A notable example being the transition to weekly reporting for incidence of COVID-19 in the majority of countries. This change in the regularity of reporting reduces the burden on the health system, but provides new challenges to any inference techniques developed to understand transmission. To test the reduced data model, we aggregate our data into randomly varying windows. The choice of random windows was made as we expect that this is a worst case scenario and captures all arbitrary reasons for data aggregation. In Figure 5 we see that the application of the reduced data model to randomly aggregated data still captures the majority of trends in the effective reproduction number and daily incidence. We conclude that our model is robust to arbitrary changes in window size.

**Fig 5.**
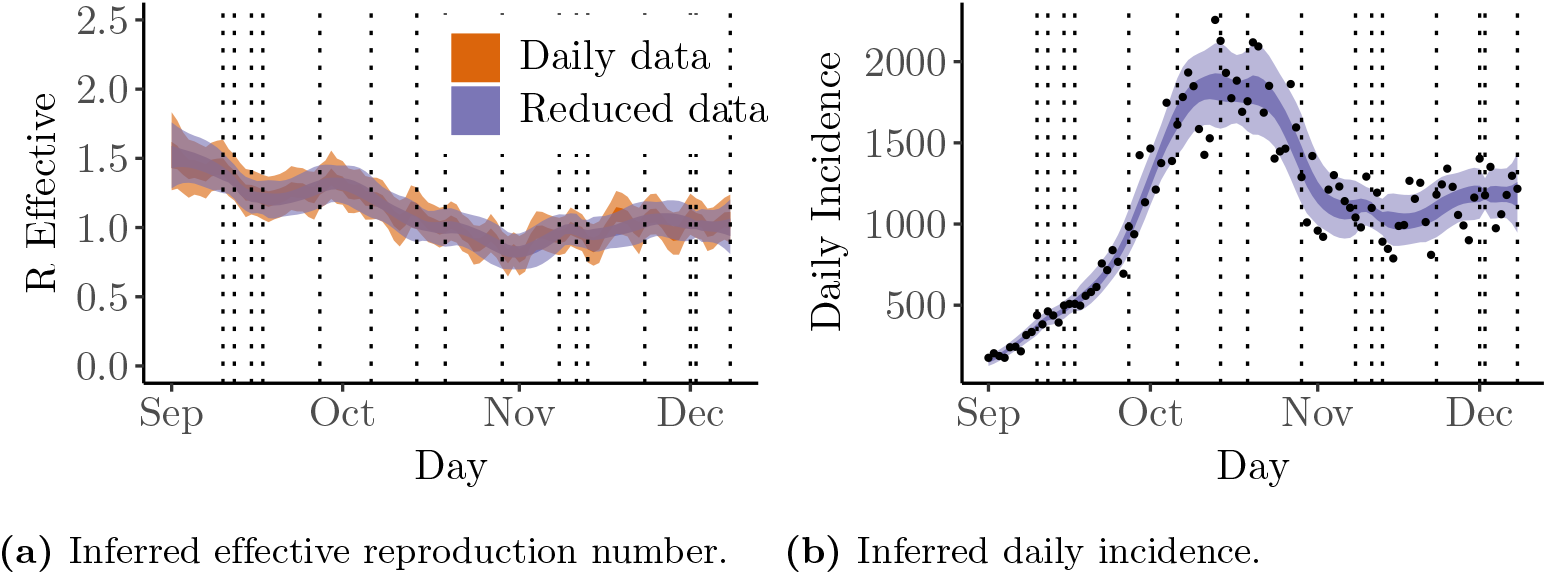
Inferring the effective reproduction number and daily incidence from randomly aggregated windows of real data from the COVID-19 pandemic in Melbourne, Victoria. Here, *α* = *β* = 10^−3^ and *λ* = log 2. The ribbons correspond to the 50% and 95% quantiles. The dashed lines are the day of reporting.

## Discussion

The COVID-19 pandemic drew considerable attention to the importance of infectious disease surveillance, however, in some regions such surveillance can be hindered by lack of funding and and weak health systems. Notably, surveillance of infectious diseases transmission within lower and middle income countries often suffer from irregular reporting and data dropout whereas high income countries will change their reporting windows depending upon governmental priorities. Therefore, there is a need to develop tools that are able to efficiently and flexibly handle changes to disease surveillance data streams, including aggregation and unforeseen dropout. In this work we provide a novel inference method that can efficiently handle data aggregation and dropout within a single framework. While the results here have focused on cases of COVID-19 reported in Victoria, Australia, the technique is trivially applicable to any other region and a wide variety of notifiable infectious diseases.

The results presented in this work demonstrate that we can accurately infer both the effective reproduction number and daily incidence even with 50% of observations missing. Data dropout is problematic in all regions, as managing data streams from multiple sources is not trivial and is subject to failure at a moments notice, but is especially problematic in low and middle income regions. In developing the reduced data model we remove the requirement of consistent measurements, instead placing a prior of a widely used infection model over our daily incidence. The benefit of this approach is that it allows us to apply the reduced data model without having to resort to *a-priori* imputation or other approximations for any missing or aggregated data. Across all examples that we consider, with both simulated and real data, the reduced data model has been able to accurately infer both the effective reproduction number and the daily incidence, resulting in comparable measurements to a more standard daily data model. The reduced data model is a robust technique that can be used to help understand spread of infectious disease even in regions with limitations to the surveillance data stream.

Unfortunately, it is not always the case that disease notification systems are able to work with a consistent data window. For example, areas where communication protocols are prone to failure (i.e., lack of internet access), may not be able to report results on day of diagnosis. Due to the communication failure case numbers may either not come through at all or be reported in one aggregated dataset, missing the information of when each diagnosis occurred. These arbitrary changes to the reporting window does not effect the application of the reduced data model described in this work. In such circumstances, regions could augment their data source with the known window of communication failure (a simple measurement to track and report with case data) and still be able to accurately determine the effective reproduction number. In Figure 5 and Appendix A.4 we provide notable examples where the reduced data model estimates both the daily incidence and effective reproduction number with great accuracy on data sets with random aggregation windows. This great flexibility exhibited by the reduced data model may prove invaluable to health systems as unforeseen challenges will not stall surveillance.

Another common example of where data aggregation may be beneficial is that of weekend effects, where case reporting is delayed over the weekend due to lower staff counts, resulting in inflated counts at the start of the week. It may be suitable to use aggregated data over the weekend to smooth out the inflated case counts. The performance of the reduced data model with random aggregation windows highlights the applicability of our technique for any arbitrary reason of data aggregation.

A further benefit to the reduced data model is that inference times across all examples provided are extremely quick, taking seconds on a laptop. As such we have not explored the application of other techniques such as particle filtering [21, 22], which is beneficial for sequential measured data. We argue that these approaches are not required as they would introduce extra problems such as particle degeneracy. In comparison, the Hamiltonian Monte Carlo method used within Stan encodes all information in the posterior and does not suffer from information loss during sampling. Furthermore, the speed at which inferences can be made make this tool suitable for use within any country, not requiring the use of large computational clusters.

A limitation of this tool is that it is in an early stage of development in comparison to the daily techniques described previously in the literature [ 2–6.] These tools have had significant development throughout the COVID-19 pandemic to ensure that they were inferring appropriate statistics on the effective reproduction number. However, there was minimal focus on working with reduced data sources such as what we are facing now. Whilst our approach does not yet incorporate delays and right truncation, as raised in [23], our work does address the shortcomings present in other tools when dealing with imperfect data such as incomplete observations and aggregation. In future work, we can focus on incorporating suggestions highlighted in [23].

A notable exclusion from our algorithm is that we do not explicitly account for uncertainty in the generation interval. This uncertainty is an important consideration as the effective reproduction number is sensitive to the choice of generation interval [23, 24]. We note that it is rather trivial to extend the posterior and make the generation interval a random variable as well, however for the work investigated here we decided against incorporating this added complexity at this stage. As such we recommend that researchers be aware of this shortcoming and test their results with varying generation intervals as done by [11].

To leverage the extremely efficient Hamiltonian Monte Carlo sampling algorithm within Stan [17], we use a normal approximation for the Poisson distribution for the daily incidence random variables. This approximation is only valid when daily incidence numbers are away from zero. This has not been a problem in all cases considered in this work, however, if this work was to be applied to a near elimination scenario, this assumption may break down. In near elimination settings we argue that the use of this method is impractical, as the case numbers are so small that there would be minimal benefit to aggregated reporting and the full line list of infection should be tracked instead. For such settings, it may be more suitable to consider a case specific reproduction number as has been previously investigated with malaria transmission [25]. If elimination settings are to be considered within this framework, in the limit of small incidence it is possible to marginalise out the inferred daily incidence and to use Stan on the marginalised posterior for effective reproduction number.

Finally we highlight an interesting observation made during our analysis. It appears that there is a trade off in measuring over-dispersion and the effective reproduction number. We have set *α* = *β* =1 × 10^*-*3^ for all results within the main text, ensuring that the prior on *σ* _*R*_ is uninformative. However, we have noticed that the Bayesian missing information criterion is not satisfied when the data set becomes extremely spare, namely, a data set that consists of 50% dropout and weekly aggregation. During investigation of this extremely sparse data set, it appears that there is a complicated interaction between measuring the over-dispersion parameter and daily fluctuations of the effective reproduction number. If the variance of the random walk is not constrained then the over-dispersion parameter is not identifiable. As such, when dealing with an extremely sparse data set, we hypothesise that the prior on *σ* _*R*_ must become more informative or the ability to infer the over-dispersion parameter is decreased. The importance of the over-dispersion parameter is arguable and the use of a Poisson distribution instead of a Negative Binomial should remove this limitation. With our use of a penalised complexity prior, we also investigated *λ* = − log(*A*) where *A* ∈ {0.3, 0.4, 0.5, 0.6, 0.7} and find that the posterior estimates are insensitive to the prior distribution specified on *ϕ*, see Figure C.1.

## Conclusion

There is a clear need for robust and efficient inference methods for infectious disease surveillance that can handle missing and aggregated data. In this work, we effectively address this need by developing a novel mathematical model that can simply and efficiently infer both the daily incidence and effective reproduction number. We have conclusively demonstrated the applicability of our approach both on real and simulated data sets. Our method is robust to random windows of aggregation, overlap and data dropout, making it applicable to regions with limited health care capacity as well as those with large amounts of resources but have decided to reduce the reporting window.

## Data Availability

All data produced in the present study are available upon reasonable request to the authors

## Footnotes

1 NegBin 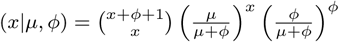

