## Supplementary Material for "Joint estimation of the effective reproduction number and daily incidence in the presence of aggregated and missing data"

### A Simulated data experiments

#### A.1 Effect of aggregation on shocks

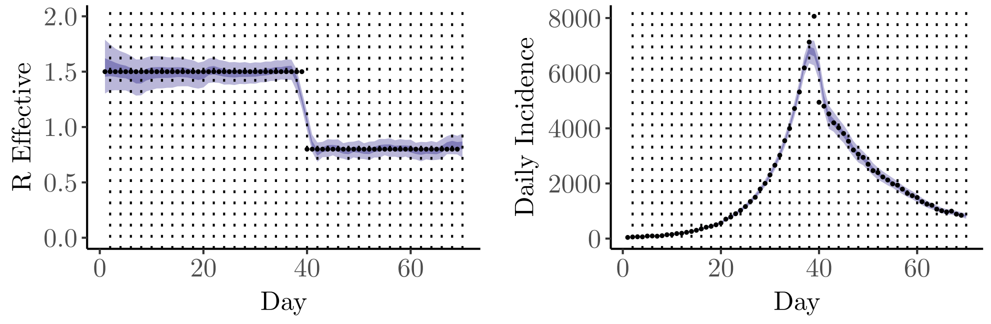


**(a)** Inferred effective reproduction number. **(b)** Inferred daily incidence.

**Fig A.1.** Application of the proposed inference method to two day aggregated simulated data. The ribbons correspond to the 50% and 95% quantiles. The dashed lines indicate the aggregation windows.

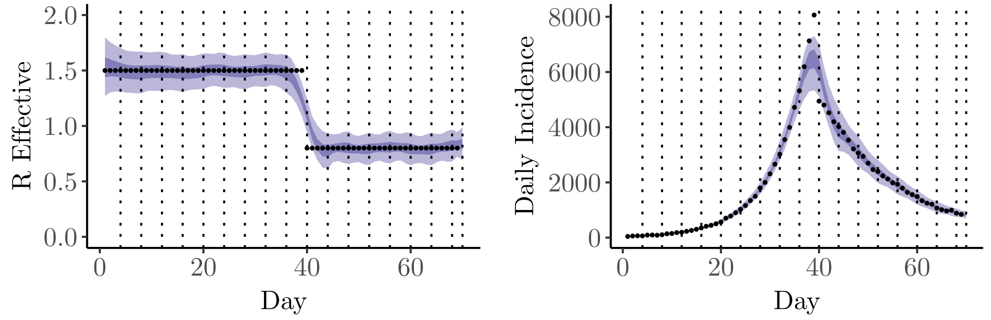


**(a)** Inferred effective reproduction number. **(b)** Inferred daily incidence.

**Fig A.2.** Application of the proposed inference method to four day aggregated simulated data. The ribbons correspond to the 50% and 95% quantiles. The dashed lines indicate the aggregation windows.

#### A.2 Aggregation on repeated waves

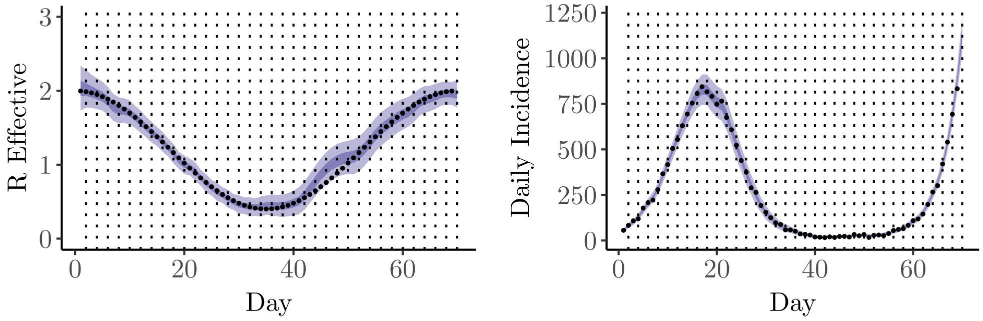


**(a)** Inferred effective reproduction number. **(b)** Inferred daily incidence.

**Fig A.3.** Application of the proposed inference method to two day aggregated simulated data. The ribbons correspond to the 50% and 95% quantiles. The dashed lines indicate the day of reporting.

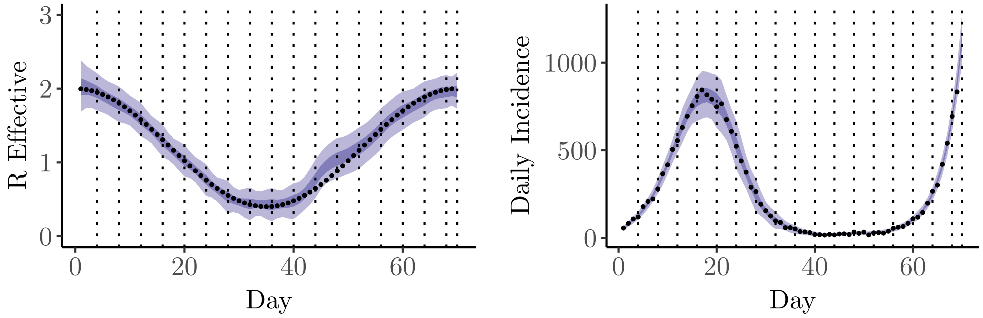


**(a)** Inferred effective reproduction number. **(b)** Inferred daily incidence.

**Fig A.4.** Application of the proposed inference method to four day aggregated simulated data. The ribbons correspond to the 50% and 95% quantiles. The dashed lines indicate the day of reporting.

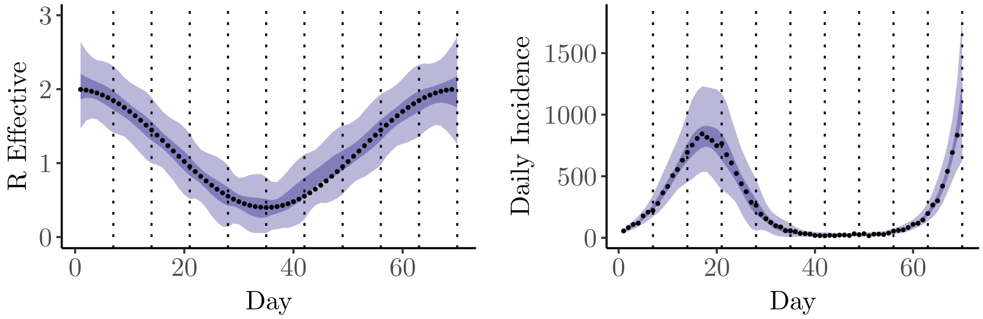


**(a)** Inferred effective reproduction number. **(b)** Inferred daily incidence.

**Fig A.5.** Application of the proposed inference method to seven day aggregated simulated data. The ribbons correspond to the 50% and 95% quantiles. The dashed lines indicate the day of reporting.

#### A.3 Dropout and aggregation

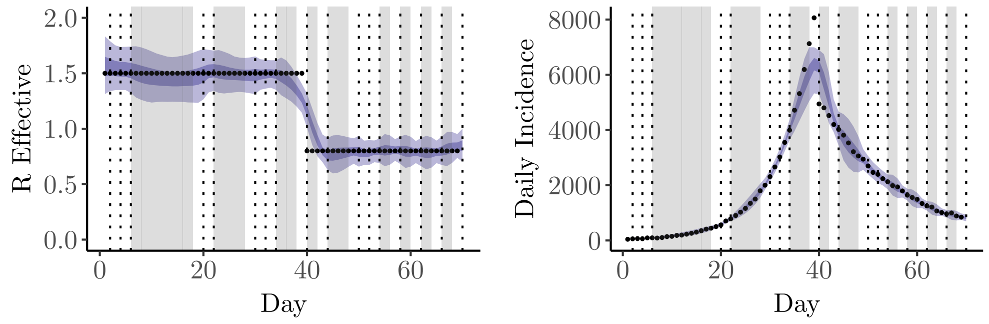


**(a)** Inferred effective reproduction number. **(b)** Inferred daily incidence.

**Fig A.6.** Application of the proposed inference method to two day aggregated simulated data with 50% drop out. The ribbons correspond to the 50% and 95% quantiles. The dashed lines indicate the aggregation windows and the grey areas indicate data dropout.

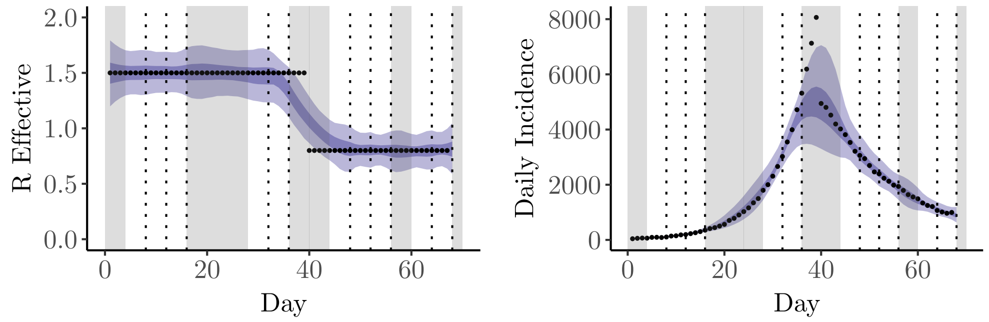


**(a)** Inferred effective reproduction number. **(b)** Inferred daily incidence.

**Fig A.7.** Application of the proposed inference method to four day aggregated simulated data with 50% drop out. The ribbons correspond to the 50% and 95% quantiles. The dashed lines indicate the day of reporting and the grey areas indicate data dropout.

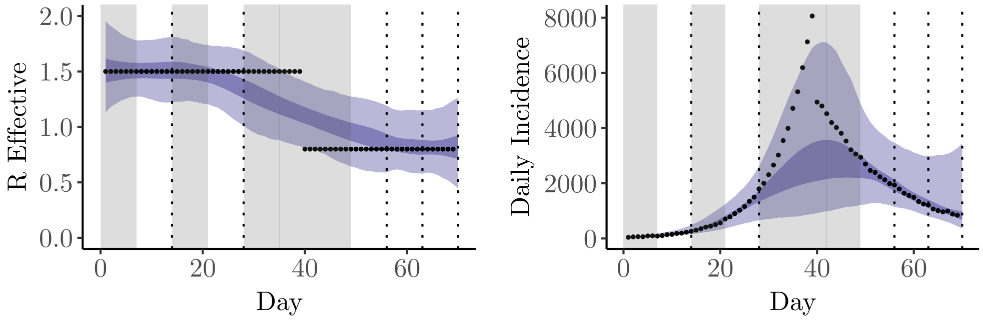


**(a)** Inferred effective reproduction number. **(b)** Inferred daily incidence.

**Fig A.8.** Application of the proposed inference method to seven day aggregated simulated data with 50% drop out. The ribbons correspond to the 50% and 95% quantiles. The dashed lines indicate the day of reporting and the grey areas indicate data dropout.

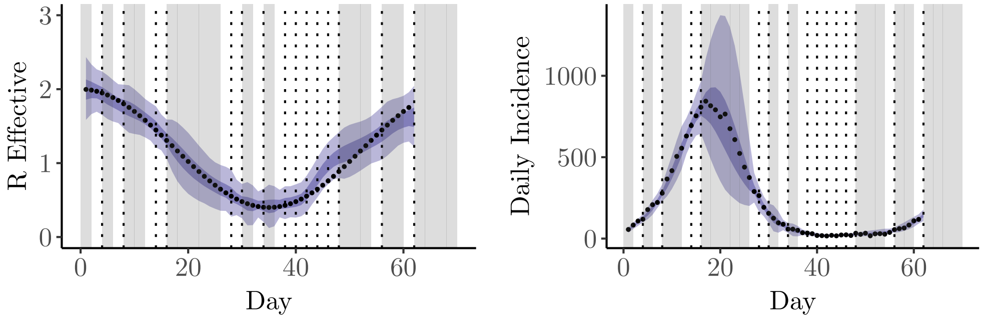


**(a)** Inferred effective reproduction number. **(b)** Inferred daily incidence.

**Fig A.9.** Application of the proposed inference method to two day aggregated simulated data with 50% drop out. The ribbons correspond to the 50% and 95% quantiles. The dashed lines indicate the day of reporting and the grey areas are regions of incomplete data.

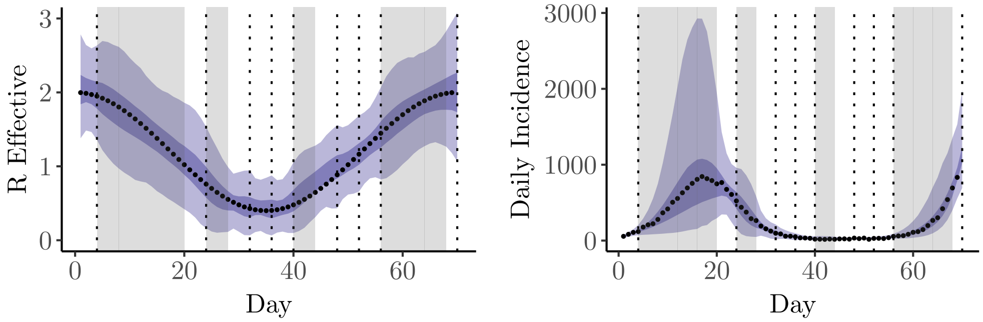


**(a)** Inferred effective reproduction number. **(b)** Inferred daily incidence.

**Fig A.10.** Application of the proposed inference method to four day aggregated simulated data with 50% drop out. The ribbons correspond to the 50% and 95% quantiles. The dashed lines indicate the day of reporting and the grey areas are regions of incomplete data.

#### A.4 Random aggregation windows

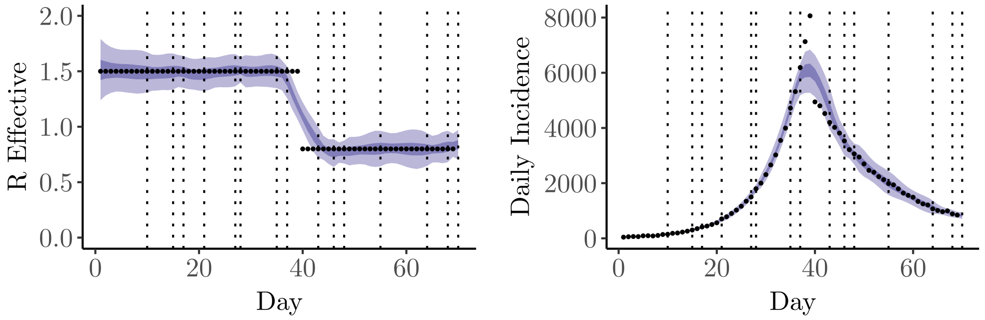


**(a)** Inferred effective reproduction number. **(b)** Inferred daily incidence.

**Fig A.11.** Application of the proposed inference method to randomly aggregated simulated data. The ribbons correspond to the 50% and 95% quantiles. The dashed lines indicate the day of reporting.

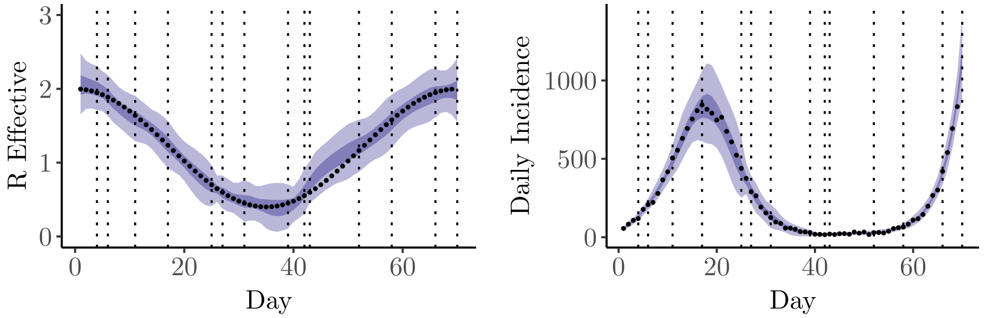


**(a)** Inferred effective reproduction number. **(b)** Inferred daily incidence.

**Fig A.12.** Application of the proposed inference method to randomly aggregated simulated data. The ribbons correspond to the 50% and 95% quantiles. The dashed lines indicate the day of reporting.

### B Real data dropout and aggregation

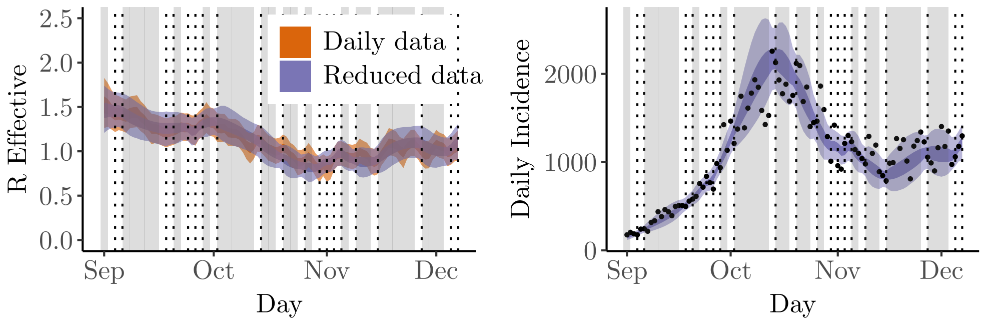


**(a)** Inferred effective reproduction number. **(b)** Inferred daily incidence.

**Fig B.1.** Results for a two day aggregation window with 50% dropout. The ribbons correspond to the 50% and 95% quantiles. The dashed lines indicate the day of reporting and the grey areas are regions of incomplete data.

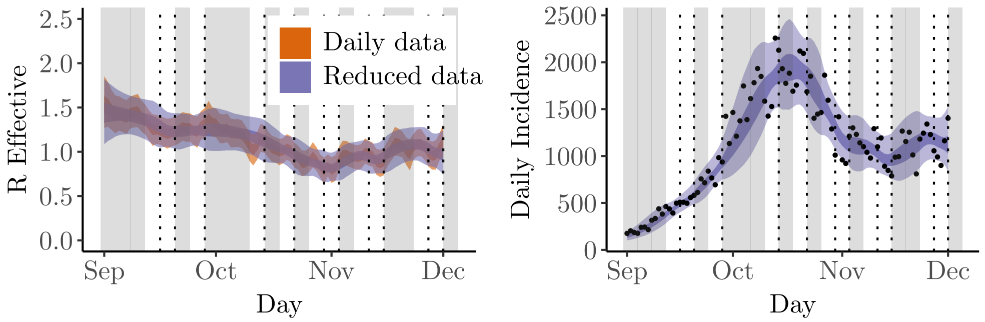


**(a)** Inferred effective reproduction number. **(b)** Inferred daily incidence.

**Fig B.2.** Results for a four day aggregation window with 50% dropout. The ribbons correspond to the 50% and 95% quantiles. The dashed lines indicate the day of reporting and the grey areas are regions of incomplete data.

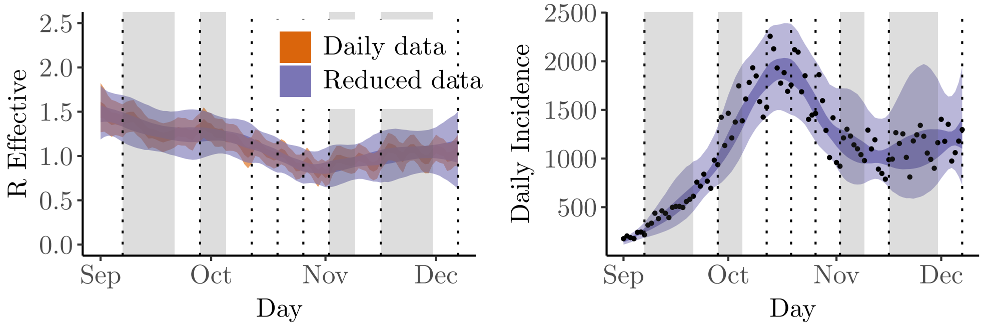


**(a)** Inferred effective reproduction number. **(b)** Inferred daily incidence.

**Fig B.3.** Results for a seven day aggregation window with 50% dropout. The ribbons correspond to the 50% and 95% quantiles. The dashed lines indicate the day of reporting and the grey areas are regions of incomplete data.

#### C Choice of penalised complexity prior

The choice of length scale in the penalised complexity prior can be recast into understanding the prior density we desire to be in the tails of the distribution. To do so, we use a transformed variable $\alpha = 1/\phi$, this way, $\alpha = 0$ corresponds to a Poisson distribution and increasing $\alpha$ increases the over-dispersion in the system. The corresponding penalised complexity prior on $\alpha$ is,

$$p(\alpha) = \frac{\lambda}{2\sqrt{\alpha}} e^{-\lambda\sqrt{\alpha}}.$$

To define the value of $\lambda$, we determine how much probability density, $A$, we desire in our prior distribution above a threshold $U$. That is, we choose $\lambda$ such that,

$$p(\alpha > U) = e^{-\lambda\sqrt{U}} = A,$$

which has a solution

$$\lambda = -\frac{\log(A)}{\sqrt{U}}.$$

In our results we want to the majority of the density to be between zero and one as we want to assume that it is Poisson distributed in its simplest form. We present our results with multiple different values of $A$, corresponding to 50%, 60% and 70% of prior density lying in the range $\alpha > 1$. In Figure C.1 we show the results of our estimates across $A \in \{0.3, 0.4, 0.5, 0.6, 0.7\}$.

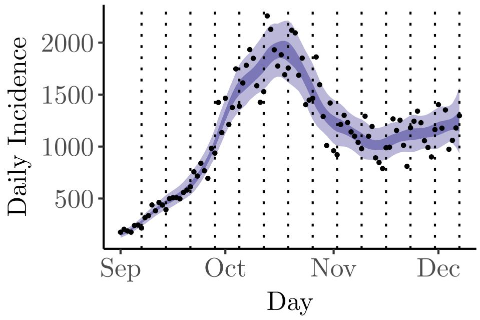


**(a)** $A = 0.3$.

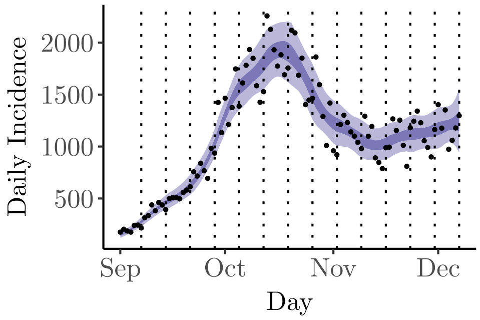


**(b)** $A = 0.4$.

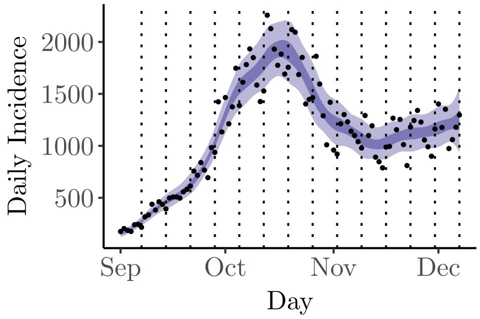


**(c)** $A = 0.5$.

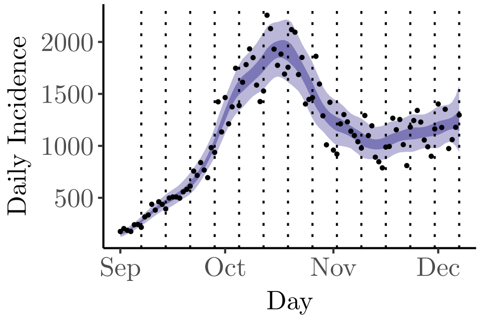


**(d)** $A = 0.6$.

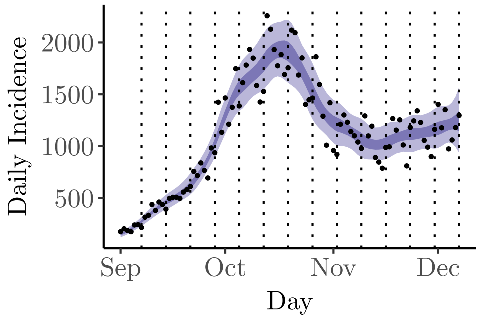


**(e)** $A = 0.7$.

**Fig C.1.** Here we show results for seven day windows of aggregation across multiple values of $\lambda$. The results shown are independent of the choice of $\lambda$.
